# Respiratory Syncytial Virus burden among Ugandan Adults Aged ≥65 Years: A 15-Year Sentinel Surveillance Study of Prevalence, Coinfections, and Comorbidities (2010–2025)

**DOI:** 10.1101/2025.09.15.25335786

**Authors:** Haruna Muwonge, Joyce Namulondo, Levicatus Mugenyi, Joweria Nakaseegu, Bridget Nakamoga, Esther Amwine, Roselyne Akugizibwe, Abdul Ssekandi, Mustafa Ssaka, Hassan Kasujja, Godfrey S Bbosa, David Odongo, Julius Lutwaama, John Kayiwa, Bruce Kirenga, Barnabas Bakamutumaho

## Abstract

**Background:** Respiratory syncytial virus (RSV) is an important cause of acute respiratory illness in older adults, yet data from sub-Saharan Africa remain scarce. Understanding prevalence, coinfections, and risk factors in this age group is critical for guiding surveillance and prevention.

**Methods:** We conducted a retrospective cross-sectional study using Uganda’s national influenza-like illness (ILI) and severe acute respiratory infection (SARI) sentinel surveillance data from December 2010 to January 2025. Adults aged ≥65 years with Real time-PCR (RT-PCR) results for RSV, influenza A/B, and SARS-CoV-2 were included. Descriptive analyses summarized period prevalence, clinical characteristics, and temporal trends. Poisson regression with robust variance estimated adjusted prevalence ratios (aPRs) for factors associated with RSV infection and hospitalization.

**Results:** Among 545 illness episodes (mean age 73.2 years; 54.1% female), the period prevalence of RSV across 2010–2025 was 4.8% (95% CI: 3.3–6.9), comparable to influenza A (4.2%) and lower than SARS-CoV-2 (6.4%). Most RSV cases were mono-infections (92.3%), with rare RSV–influenza coinfections (0.4%) and no RSV–SARS-CoV-2 coinfections. Asthma (aPR 6.08, 95% CI: 1.18–31.26, p=0.031) and pneumonia (aPR 2.83, 95% CI: 1.06–7.56, p=0.038) were independent predictors of RSV infection. Hospitalization was strongly associated with asthma (aPR 21.69, 95% CI: 7.50–62.71, p<0.001), pneumonia (aPR 3.80, 95% CI: 1.51–9.56, p=0.005), and heart disease (aPR 3.50, 95% CI: 1.03–11.91, p=0.045). RSV activity showed seasonal peaks in March and June, with a marked decline during 2020–2021 (COVID-19 restrictions) and resurgence thereafter.

**Conclusions:** RSV is a consistent contributor to respiratory illness among older Ugandan adults, with period prevalence estimates similar to influenza A and clinically important associations with asthma and pneumonia. Seasonal peaks during the dry season and post-pandemic resurgence emphasize the need for RSV integration into surveillance and the timely introduction of preventive interventions, including vaccination, in low- and middle-income settings.

## Introduction

Respiratory Syncytial Virus (RSV) is a leading cause of acute lower-respiratory infection in children and imposes a substantial, though often under-appreciated, burden among older adults worldwide. Recent estimates show that adults’ ≥65 years account for 123,000–193,000 RSV-associated hospitalisations annually in high-income countries, with the highest rates in those ≥75 years (Havers et al., 2023; Havers et al., 2024). In the United States, RSV-attributable hospitalisation rates—about 25 cases per 10,000 persons—already rival or exceed those of influenza in this age group, and nearly 60% of PCR-confirmed adult cases require inpatient care (Trifonov et al., 2025; Widmer et al., 2012). In low- and middle-income countries (LMICs), adult-specific data remain scarce, though pediatric studies highlight RSV’s impact. In 2015, an estimated 33.1 million RSV-associated ALRI episodes occurred in children <5 years, leading to ∼3.2 million hospitalisations and ∼118,200 deaths (including ∼59,600 in-hospital deaths), with >95% of cases and >97% of deaths in LMICs (Li et al., 2022; Shi et al., 2017), prompting the World Health Organization to prioritise RSV in children alongside influenza (World Health Organization, 2025).

In older adults, the impact of RSV is amplified by immunosenescence, comorbidities, and coinfections. Incidence has been estimated at about 600 cases and 157 hospitalizations per 100,000 person-years among those aged ≥60, yet its true burden remains poorly defined, particularly in low-income countries such as Uganda (Kenmoe & Nair, 2024). For example, pooled data from Kenya, Tanzania, and Uganda reported an overall adult RSV prevalence of only 3%, but with substantial heterogeneity and a lack of age-stratified estimates (Therese Umuhoza et al., 2021).

Since 2010, Uganda’s Ministry of Health implemented a nationwide health facility-based sentinel surveillance for influenza-like illness (ILI) and severe acute respiratory infection (SARI), incorporating PCR testing for multiple respiratory viruses and, more recently, integrating SARS-CoV-2 diagnostics (Kayiwa et al., 2023). To date, analyses have primarily focused on pediatric cohort or influenza trends, leaving older adults largely understudied. Understanding RSV epidemiology in this population is especially urgent, as Uganda’s population aged ≥60 years is projected to double by 2050, increasing the demand for geriatric respiratory care. Hence, this study’s 15-year surveillance window (2010-2025) spanning the pre- and post-COVID-19 eras offers a unique opportunity to characterise RSV burden, including seasonality. This is particularly relevant given the increasing availability of approved RSV vaccines for adults, such as Arexvy (GSK) and Abrysvo (Pfizer), both of which received FDA regulatory approval in 2023 (European Medicines Agency, 2023; U. S. FDA & Administration, 2023), Against this backdrop, we conducted a retrospective analysis of Uganda’s Ministry of Health sentinel ILI/SARI data to estimate RSV prevalence among adults aged ≥ 65 years, describe coinfection patterns with influenza A/B and SARS-CoV-2, investigate associations between RSV positivity with presenting symptoms including selected comorbidities and hospitalization, and examine temporal trends spanning 2010–2025.

## Methods

### Study design

We conducted a retrospective cross-sectional study using routinely collected health facility-based data from Uganda’s national influenza-like illness (ILI) and severe acute respiratory infection (SARI) sentinel surveillance system. The analysis covered the period from December 1, 2010, through January 31, 2025.

### Surveillance network and study setting

The national sentinel surveillance network comprises 16 public and private healthcare facilities across 11 districts in Uganda, representing diverse ecological and demographic contexts including urban, peri-urban, and rural areas. Facilities included regional referral hospitals, general hospitals, and health centers at levels III and IV. Sites were selected to ensure geographical representativeness. Surveillance involved routine enrolment of outpatients meeting the WHO definition for ILI (an acute respiratory infection with measured fever of ≥38 °C and cough, with onset within the last 10 days) and hospitalized patients meeting SARI criteria (an acute respiratory infection with a history of fever or measured fever of ≥38 °C and cough, with onset within the last 10 days, requiring hospitalization). Clinical and laboratory data from participating facilities were centrally collated at the Uganda Virus Research Institute (UVRI) National Influenza Centre (NIC), a WHO-accredited influenza laboratory.

### Study population and eligibility criteria

The study population consisted of adults aged 65 years or older who presented to sentinel facilities with symptoms fulfilling WHO-defined criteria for ILI or SARI and who had nasopharyngeal or oropharyngeal specimens tested for respiratory pathogens, including RSV, Influenza viruses sub-type A and B, and SARS-CoV-2. We excluded individuals with missing demographic or laboratory data, repeat visits within a 14-day window for the same illness, individuals vaccinated against RSV or influenza during the study period, those with severe immunosuppressive conditions, and patients presenting with respiratory symptoms due to non-infectious etiologies.

### Data collection and study variables

Clinical and demographic data collected included age, sex, clinical symptoms (e.g., cough, fever, sore throat, shortness of breath, fatigue, and diarrhoea), underlying comorbidities (hypertension, diabetes, heart disease, HIV infection, asthma, active or prior tuberculosis, and cancer), and hospitalization status. Laboratory data were matched to clinical records using unique patient identifiers. The primary study outcome was laboratory-confirmed RSV infection by real-time reverse transcription polymerase chain reaction (RT-PCR). Secondary outcomes included coinfection with Influenza sub-types A and B and SARS-CoV-2, seasonal trends, and associations between RSV positivity and symptoms and sverity or comorbid conditions.

### Laboratory procedures

Nasopharyngeal and/or oropharyngeal swabs were collected using sterile Dacron swabs, placed in viral transport medium, and stored at 4°C at sentinel facilities. Samples were shipped bi-weekly to UVRI-NIC under standardized cold-chain conditions. At UVRI-NIC, total nucleic acid was extracted using the Andis Viral RNA Extraction Kit. RSV detection was performed using the Healgen RSV real-time RT-PCR assay, and influenza A/B and SARS-CoV-2 were detected using the CDC Flu SC2 multiplex RT-PCR assay run on the ABI 7500 PCR platform. Samples with cycle threshold (Ct) values below 40 for RSV and SARS-CoV-2, and below 36 for influenza viruses, were considered positive. Quality control measures included extraction controls, positive and negative amplification controls, and human RNase P internal controls in all test runs. UVRI-NIC participates annually in WHO external quality assessment programmes and maintains ISO-accredited laboratory practices.

### Data management

All surveillance data were anonymized, and securely stored in a password-protected database accessible only by authorized investigators. Data quality assurance included routine checks for missing, duplicate, or inconsistent entries and random verification against original case investigation forms. Data management adhered to the requirements of Uganda’s Data Protection and Privacy Act (Republic of Uganda, 2019).

### Statistical analysis

All analyses were conducted using Stata version 18 (StataCorp, College Station, TX, USA). Descriptive statistics summarized baseline characteristics of participants, with means and standard deviations (SD) reported for continuous variables and frequencies with percentages for categorical variables. The prevalence of RSV was calculated as proportions with 95% confidence intervals (CI). RSV prevalence was further stratified by demographic factors, clinical presentation, comorbidities, season, and surveillance period (pre-vs. post-COVID). Between-group comparisons were performed using Pearson chi-square test or Fisher’s exact test where appropriate. To identify factors associated with RSV positivity, prevalence ratios (PRs) and 95% CIs were estimated using Poisson regression model with robust variance. Multivariable model was fitted adjusting for demographic, clinical, and comorbidity variables that indicated potential association with RSV restricted to participants with complete data. Seasonal dynamics were assessed by plotting monthly positivity rates for RSV and comparator viruses, and by comparing aggregate prevalence between wet and dry rainfall periods. Temporal trends were described across the pre-pandemic, pandemic, and post-pandemic phases of surveillance. For severity analyses, hospitalization was used as a proxy outcome for severe RSV illness. Associations between comorbidities and hospitalization among RSV-positive participants were examined using Poisson regression with robust variance to estimate adjusted prevalence ratios (aPRs). A p-value <0.05 was considered statistically significant.

### Ethical considerations

Ethical approval for this retrospective analysis was obtained from the Makerere University School of Biomedical Sciences Research and Ethics Committee (SBS-REC, approval # SBS-2024-685) and the Uganda National Council for Science and Technology (UNCST, approval # HS5423ES). Given the retrospective nature of anonymized surveillance data, the SBS-REC provided a waiver of individual informed consent. The sentinel surveillance database was accessed for research purposes on July 01, 2025. The database contained only de-identified information, and authors had no access to information that could identify individual participants during or after data collection. All data handling and analyses were conducted in compliance with Uganda’s Data Protection and Privacy Act (Republic of Uganda, 2019).

## Results

### Study Cohort Profile

Between 1 December 2010 and 31 January 2025 the sentinel network recorded 545 illness episodes in adults aged ≥ 65 years that satisfied the WHO ILI/SARI case definitions and had a full respiratory-virus RT-PCR panel (RSV, Influenza sub-type A and B and SARS-CoV-2). All 545 individual records met the analytical inclusion criteria, so the final study cohort represents 100 % of age-eligible surveillance encounters during the study window (Fig 1).

**Fig 1.**
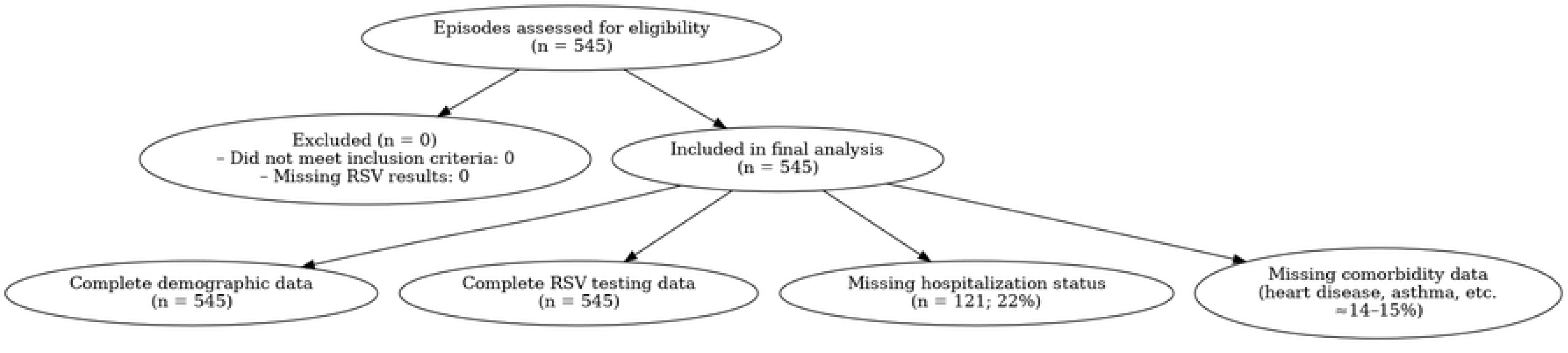
CONSORT-style flow diagram of surveillance episode inclusion and data completeness.

Completeness of the core exposure and outcome variables was high: viral PCR results, age, sex and reporting month were available for every participant. Most symptom fields showed <3 % missing data (e.g., cough 0.6 %, sore throat 2.4 %, shortness of breath 0.4 %). In contrast, several comorbidity variables were absent in 14–15 % of records (heart disease, hypertension, diabetes, asthma, active and prior tuberculosis, cancer). Hospitalization status was the single variable with substantial missingness, absent in 22.2 % of cases, while history of fever was missing in 11.4 %. Thus, missing data were largely confined to secondary clinical covariates and are unlikely to bias virological prevalence estimates.

### Baseline Characteristics of Participants

This cohort comprised 545 illness episodes in adults aged ≥ 65 years. The mean age was 73.2 ± 9.1 years, and women constituted a slight majority (54.1 %). Current smoking was uncommon (2.0 %). Cough (73.9 %), fever or history of fever (71.4 %), and shortness of breath (47.5 %) were the most frequently reported symptoms, whereas sore throat was present in 27.7 % of cases. Documented comorbidities were infrequent but clinically relevant: hypertension (9.5 %), diabetes (5.9 %), heart disease (3.9 %), and HIV infection (3.1 %); asthma, active tuberculosis, prior tuberculosis, and cancer each affected < 2 % of participants. Nearly two-thirds of encounters (65.7 %) resulted in hospital admission, although length-of-stay data were not systematically recorded. Annual enrolment counts increased sharply after 2020, reflecting the scale-up of sentinel testing during the COVID-19 period and subsequent expansion of the surveillance network. Table 1 describes the baseline characteristics of study participants.

**Table 1.**
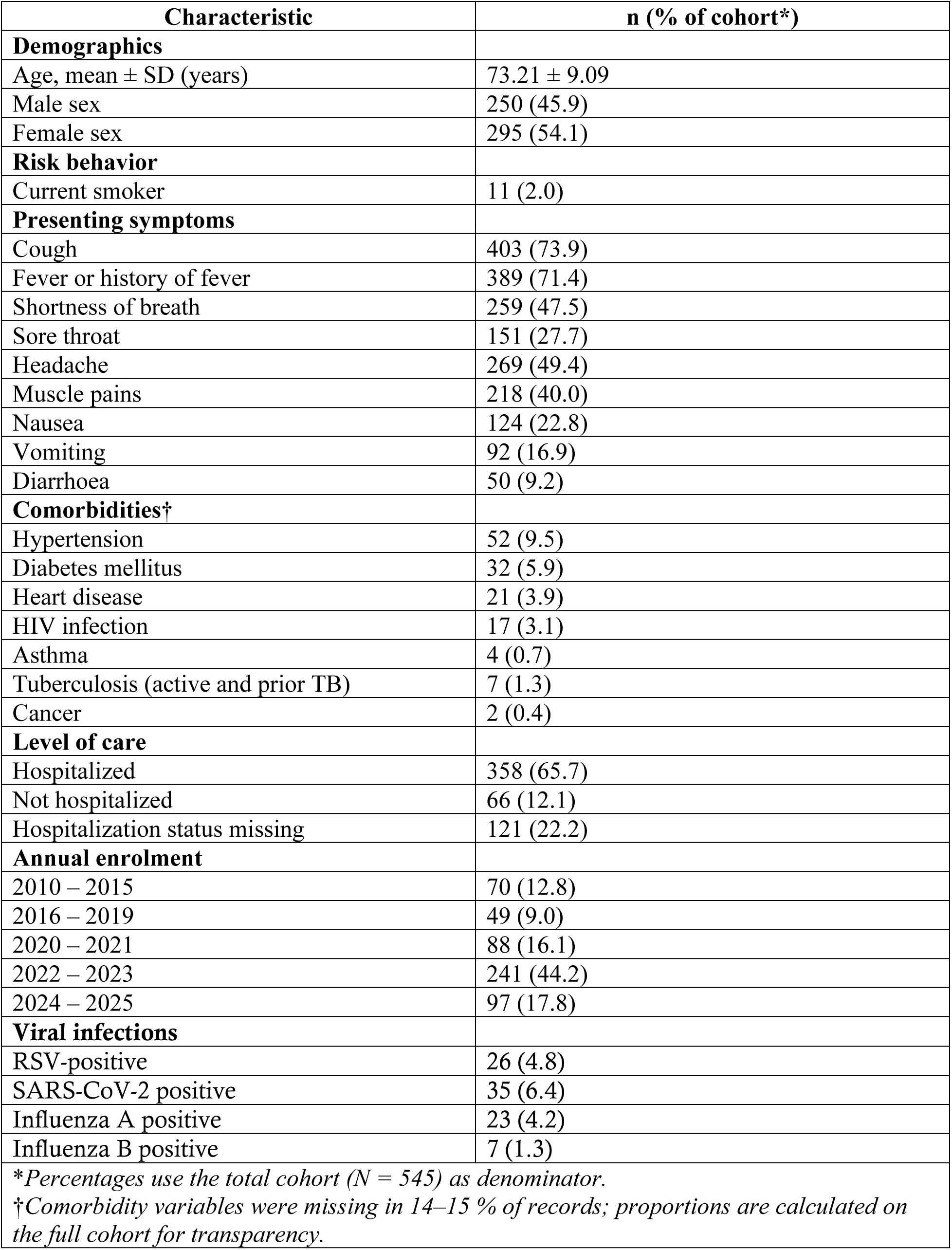
Baseline characteristics of study participants.

### Prevalence of RSV and Other Respiratory Viruses

Over the 15-year surveillance period, 26 of 545 illness episodes tested positive for RSV, yielding an overall period prevalence of 4.8% (95% CI: 3.3–6.9). Fig 2 shows the geographical distribution of confirmed RSV cases across several districts in Uganda. For comparison, the observed period prevalence was 6.4% for SARS-CoV-2, 4.2% for influenza sub-type A, and 1.3% for Influenza sub-type B (Table 1). Table 2 provides a detailed breakdown of RSV prevalence stratified by demographic group, clinical presentation, and comorbidities.

**Fig 2.**
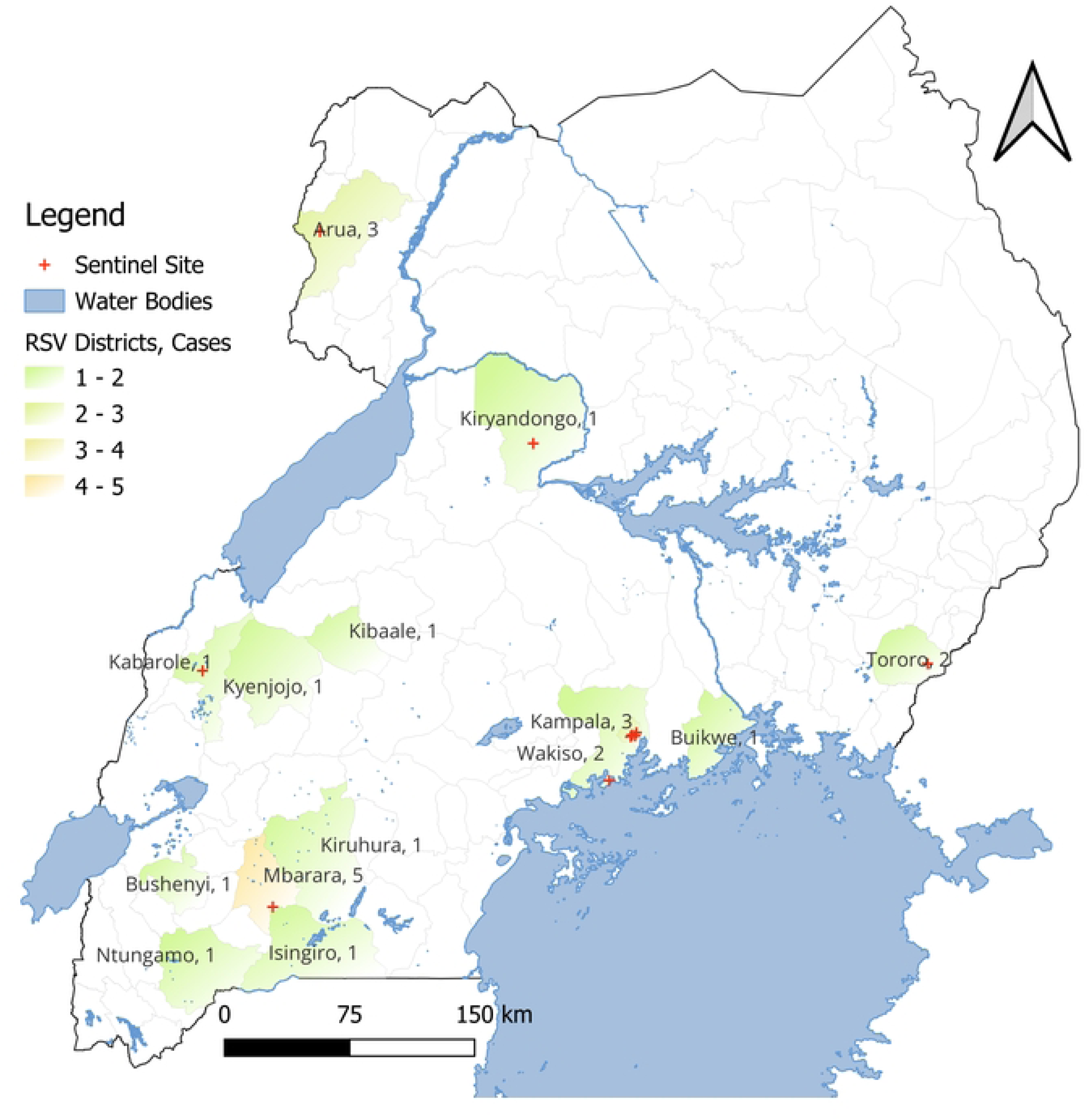
Geographical distribution of RSV-positive cases among adults aged ≥65 years in Uganda, 2010–2025.

**Table 2.** RSV prevalence by demographic, clinical, and comorbidity characteristics among adults aged ≥65 years.

| Characteristic | RSV-positive n/N (%) | 95% CI | p-value* |
| --- | --- | --- | --- |
| **Sex** |  |  |  |
| Male | 9/250 (3.6) | 1.9 – 6.8 | 0.238 |
| Female | 17/295 (5.8) | 3.6 – 9.1 |  |
| **COVID period** |  |  |  |
| Pre-COVID | 8/113 (7.1) | 3.6 – 13.8 | 0.196 |
| Post-COVID | 18/432 (4.2) | 2.6 – 6.6 |  |
| **Season** |  |  |  |
| Dry | 14/265 (5.3) | 3.2 – 8.8 | 0.585 |
| Wet | 12/280 (4.3) | 2.5 – 7.5 |  |
| **Age group** |  |  |  |
| 60–69 | 9/209 (4.3) | 2.3 – 8.2 | 0.667 |
| 70–79 | 8/189 (4.2) | 2.1 – 8.4 |  |
| 80+ | 9/147 (6.1) | 3.2 – 11.6 |  |
| **Smoking status** |  |  |  |
| Yes | 1/11 (9.1) | 1.3 – 64.5 | 0.487 |
| No | 24/521 (4.6) | 3.1 – 6.8 |  |
| **Symptoms** |  |  |  |
| Cough | 25/403 (6.2) | 4.2 – 9.1 | **0.009** |
| Sore throat | 12/151 (7.9) | 4.6 – 13.7 | **0.016** |
| Shortness of breath | 15/259 (5.8) | 3.5 – 9.5 | 0.296 |
| Fever/history of fever | 20/389 (5.1) | 3.4 – 7.9 | 0.723 |
| Vomiting | 6/92 (6.5) | 3.0 – 14.2 | 0.390 |
| Headache | 12/269 (4.5) | 2.6 – 7.8 | 0.716 |
| Nausea | 6/124 (4.8) | 2.2 – 10.6 | 0.972 |
| Muscle pains | 14/218 (6.4) | 3.9 – 10.7 | 0.144 |
| Diarrhoea | 5/50 (10.0) | 4.3 – 23.2 | 0.070 |
| **Comorbidities** |  |  |  |
| Asthma | 2/4 (50.0) | 16.1 – 1.0 | **<0.001** |
| Heart disease | 3/21 (14.3) | 4.9 – 41.8 | 0.063 |
| Pneumonia† | 6/62 (9.7) | 4.5 – 20.8 | 0.054 |
| TB (active + prior) | 1/13 (7.7) | 1.4 – 71.0 | 0.736 |
| Diabetes mellitus | 2/32 (6.2) | 1.6 – 24.4 | 0.686 |
| Hypertension | 4/52 (7.7) | 3.0 – 19.9 | 0.299 |
| HIV | 0/17 (0.0) | --- | 0.348 |
| Cancer | 0/2 (0.0) | -- | 0.736 |
| * Pearson $\chi^2$ or Fisher's exact test as appropriate. CI = confidence interval. | | | |
| † Diagnosis recorded by admitting clinician. |  |  |  |

RSV was slightly higher among women (5.8%) than men (3.6%), but the sex-based difference was not statistically significant (p = 0.238). Stratification by surveillance period showed RSV period prevalence declined from 7.1% pre-COVID (2010–2019) to 4.2% post-COVID (2020– 2025), though the difference did not reach significance (p = 0.196). RSV period prevalence was also slightly higher in the dry season (5.3%) compared to the wet season (4.3%), again with no significant difference (p = 0.585).

When examined by age, RSV period prevalence was 4.3% among adults aged 60–69, 4.2% among those 70–79, and 6.1% among those aged 80 and above (p = 0.667), showing no clear age-related significant diference. Smoking was not significantly associated with RSV (9.1% vs 4.6%, p = 0.487), though this analysis was limited by small numbers of current smokers.

Among presenting symptoms, cough and sore throat were the only significant correlates of RSV infection. Participants with cough had an RSV positivity of 6.2%, compared to 0.7% among those without cough (p = 0.009). Similarly, sore throat was reported in 7.9% of RSV-positive cases versus 3.1% of RSV-negative cases (p = 0.016). Other symptoms—including shortness of breath (5.8% vs 3.9%, p = 0.296), fever (5.1% vs 4.3%, p = 0.723), headache, nausea, vomiting, muscle pains, and diarrhoea—showed no statistically significant differences with RSV positivity.

Several comorbid conditions were explored for association with RSV positivity. Asthma emerged as the strongest predictor, with 2 of 4 individuals (50.0%) testing RSV-positive compared to 5.0% among those without asthma (p < 0.001). Heart disease (14.3% vs 4.9%, p = 0.063) and pneumonia noted by the admitting clinician (9.7% vs 4.1%, p = 0.054) modest significance. No associations were detected between RSV and diabetes mellitus, hypertension, tuberculosis (active or prior), HIV, or cancer.

### Temporal Trends and Seasonal Dynamics of RSV and Comparator Viruses

#### Annual Trends and Pandemic-Associated Disruptions

As shown in Fig 3, RSV circulation was observed in most years from 2011 to 2025, with peaks in 2012, 2016, and 2023. RSV detections dropped sharply in 2020 and 2021, coinciding with the COVID-19 pandemic and national restrictions. During the same period, SARS-CoV-2 emerged as the dominant respiratory virus, peaking at ∼17% in 2021. Influenza A activity was markedly reduced but rebounded in 2023, while influenza B showed irregular, low-level circulation, with its highest peak in 2013.

**Fig 3.**
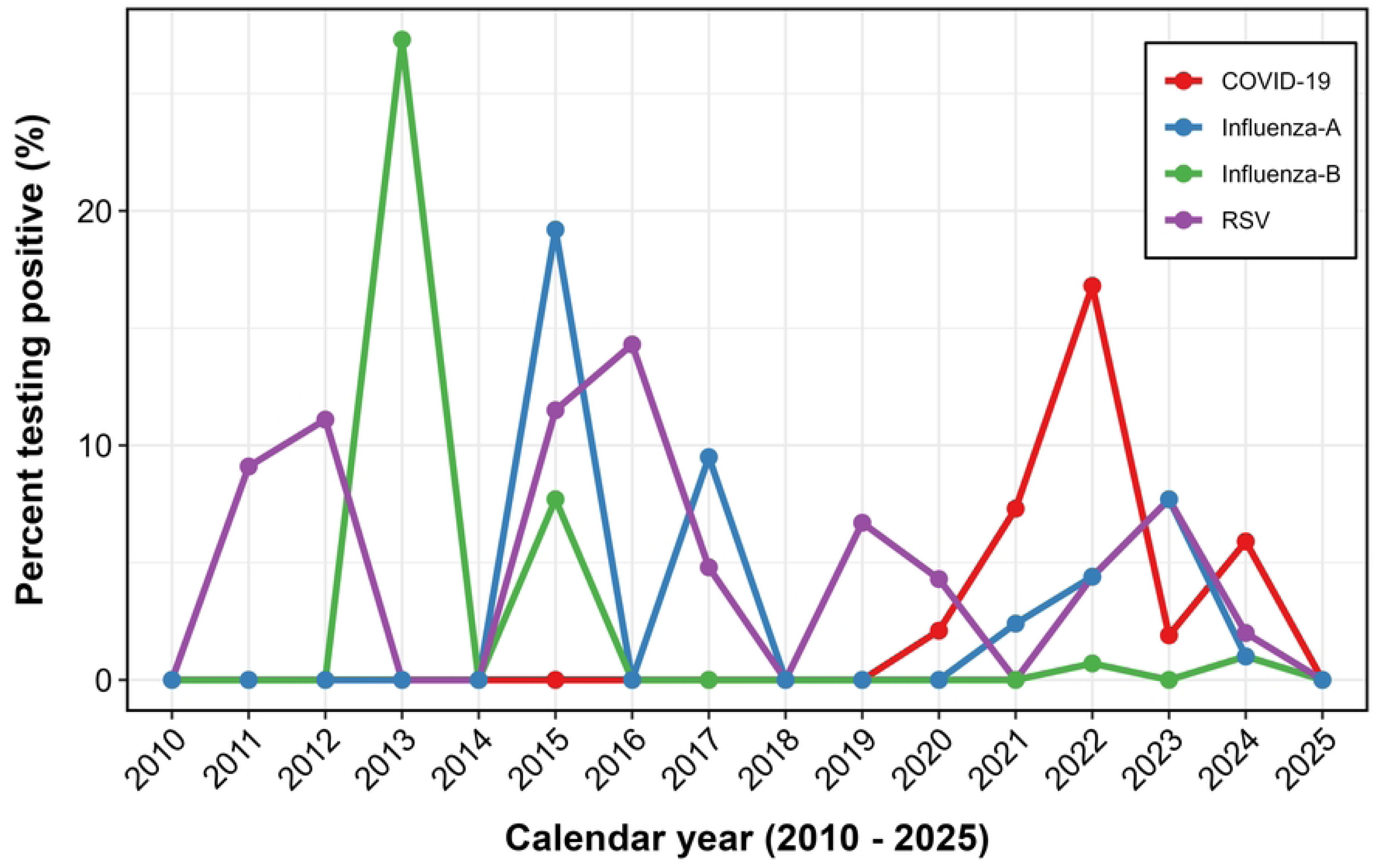
Annual positivity trends of RSV, influenza A/B, and SARS-CoV-2 among adults’ ≥65 years in Uganda, 2010–2025.

Pre- and post-pandemic stratification (Fig 4) shows RSV period prevalence declined from 7.1% before COVID-19 (2010–2019) to 4.2% afterwards (2020–2025). Influenza A and B also showed reduced activity during the pandemic, with subsequent resurgence from 2022 onwards..

**Fig 4.**
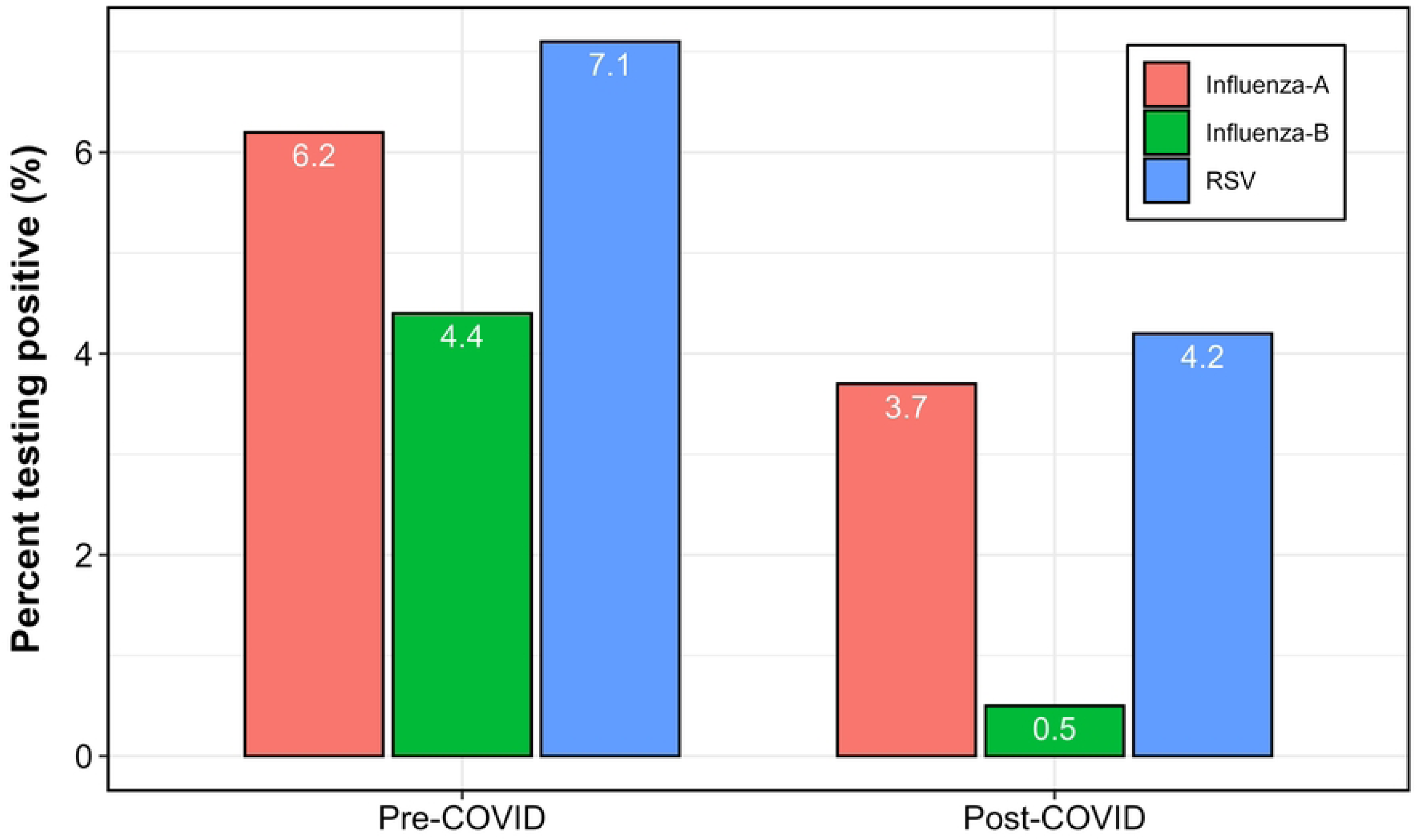
Pre- and post-COVID period prevalence of RSV, influenza A, and influenza B among adults ≥65 years in Uganda.

#### Monthly Patterns and Seasonality

Higher-resolution monthly data, shown in Fig 5, reveals the underlying seasonality of RSV, with consistent peaks in May and June; closely aligned with Uganda’s first major rainy season. In contrast, SARS-CoV-2 exhibited classic wave-like behaviour, with sharp surges in January and May–June 2021. Influenza A demonstrated less predictable seasonality, with variable peaks in March–April, while influenza B showed sporadic circulation without a discernible seasonal signature.

**Fig 5.**
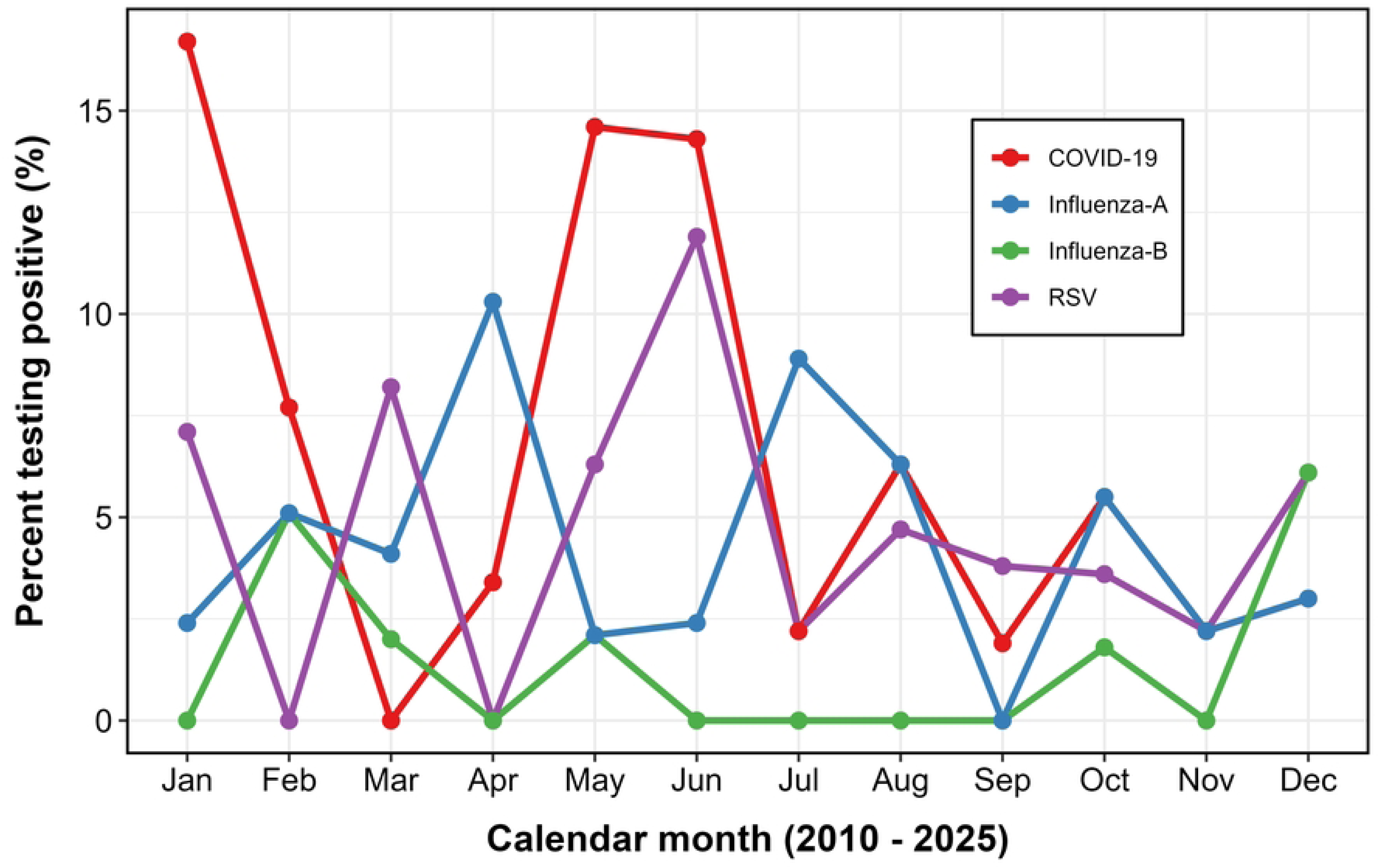
Monthly positivity trends of respiratory viruses among adults aged ≥65 years, Uganda, 2010–2025.

#### Seasonal Variation in Virus Positivity by Rainfall Period

To evaluate seasonal associations, months were classified into wet seasons (March–May and September–November) and dry seasons (June–August and December–February), following Uganda’s bimodal rainfall pattern. As illustrated in Fig 6, RSV period prevalence during dry months was 5.3%, slightly higher than the 4.3% observed in wet months. However, this difference was modest and not statistically significant (p=0.585). For comparison, SARS-CoV-2 showed a notable seasonal gradient, with dry-season positivity of 8.3 % versus 4.6 % in the wet season. This was largely driven by a pronounced peak during the January 2021 wave.

**Fig 6.**
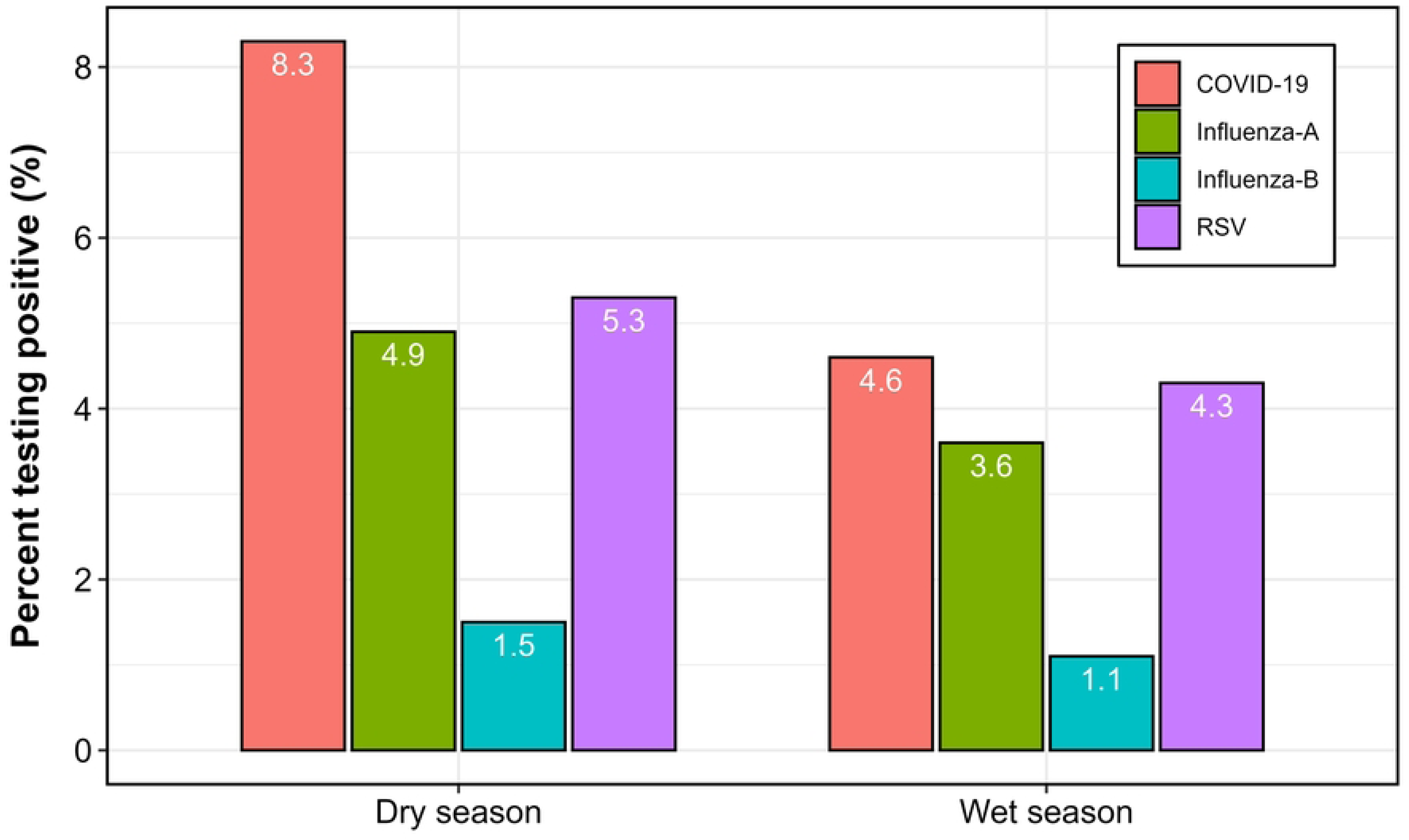
Seasonal period prevalence of respiratory viruses among adults’ ≥65 years in Uganda, 2010–2025.

Conversely, influenza A appeared more common in dry months (4.9%) than in wet months (3.6%), though the difference was small. These patterns suggest that while RSV exhibits some seasonal variation in older Ugandan adults, the association with rainfall is weaker than previously hypothesized. SARS-CoV-2 and influenza also demonstrated heterogeneous seasonal behavior, likely influenced by pandemic dynamics and public health interventions.

### RSV Multi-infection Patterns

Among the 545 illness episodes, 26 (4.8%) tested positive for RSV. Of these, 24 (92.3%) were RSV mono-infections, while 2 (0.4%) represented coinfections with influenza viruses. No instances of RSV–SARS-CoV-2 coinfection were identified. An additional 63 cases (11.6%) were positive for SARS-CoV-2 and/or influenza A or B in the absence of RSV, whereas 456 participants (83.7%) had no detectable viral pathogen.

Stratified analysis revealed several key differences across infection categories (Table 3). RSV positivity (either alone or with influenza) was more common among women (5.7%) than men (3.6%), but the difference was not statistically significant (p = 0.217). Current smokers had numerically higher rates of RSV-only infection (9.1%) and non-RSV viral infection (27.3%), but this was based on small numbers and did not reach significance (p = 0.318).

**Table 3.**
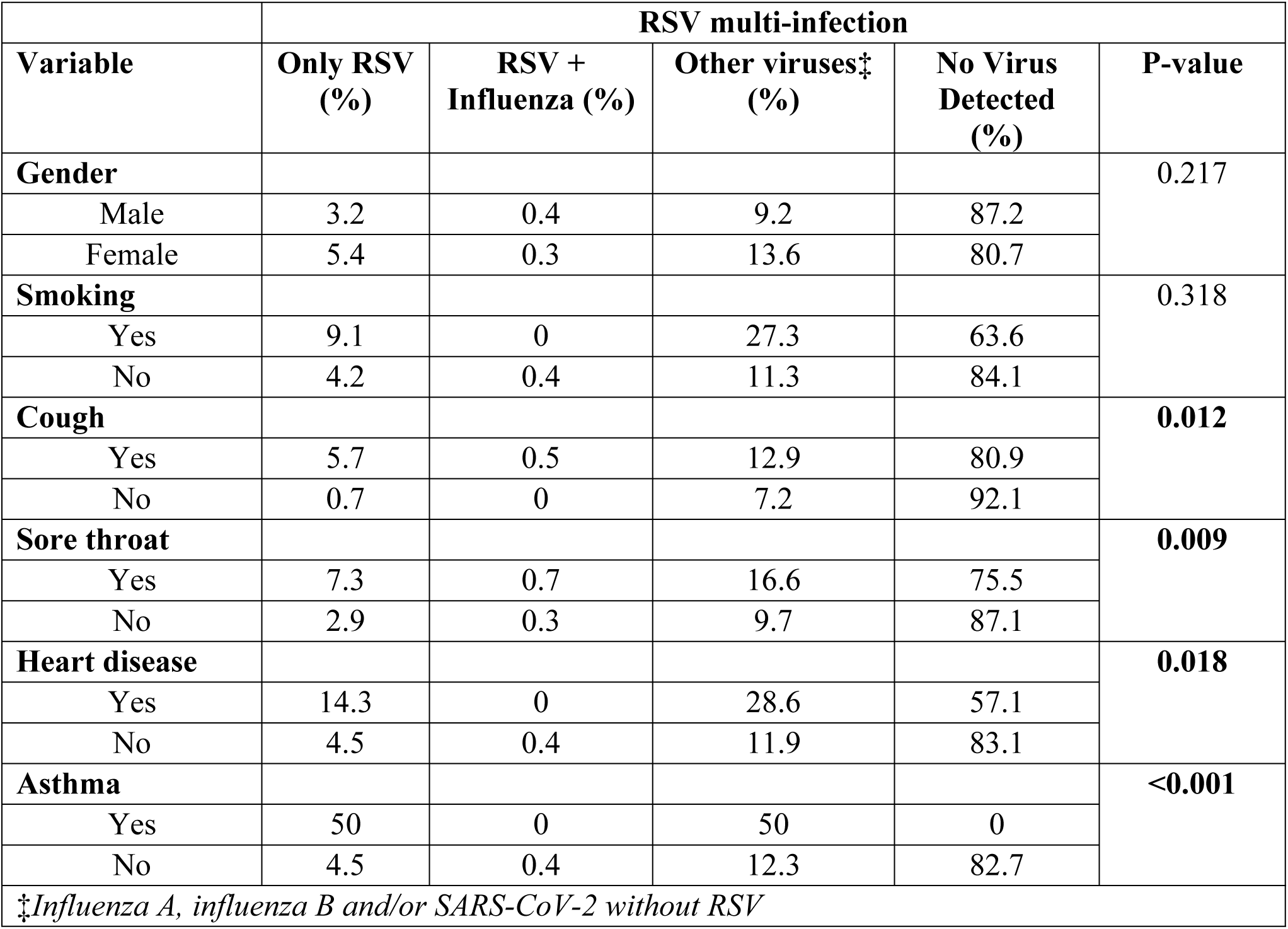
RSV mono-infection and coinfection profiles by demographic and clinical characteristics in adults aged ≥65 years.

| Variable | RSV multi-infection |  |  |  | P-value |
| --- | --- | --- | --- | --- | --- |
|  | Only RSV (%) | RSV + Influenza (%) | Other viruses‡ (%) | No Virus Detected (%) |  |
| **Gender** |  |  |  |  | 0.217 |
| Male | 3.2 | 0.4 | 9.2 | 87.2 |  |
| Female | 5.4 | 0.3 | 13.6 | 80.7 |  |
| **Smoking** |  |  |  |  | 0.318 |
| Yes | 9.1 | 0 | 27.3 | 63.6 |  |
| No | 4.2 | 0.4 | 11.3 | 84.1 |  |
| **Cough** |  |  |  |  | **0.012** |
| Yes | 5.7 | 0.5 | 12.9 | 80.9 |  |
| No | 0.7 | 0 | 7.2 | 92.1 |  |
| **Sore throat** |  |  |  |  | **0.009** |
| Yes | 7.3 | 0.7 | 16.6 | 75.5 |  |
| No | 2.9 | 0.3 | 9.7 | 87.1 |  |
| **Heart disease** |  |  |  |  | **0.018** |
| Yes | 14.3 | 0 | 28.6 | 57.1 |  |
| No | 4.5 | 0.4 | 11.9 | 83.1 |  |
| **Asthma** |  |  |  |  | **<0.001** |
| Yes | 50 | 0 | 50 | 0 |  |
| No | 4.5 | 0.4 | 12.3 | 82.7 |  |

Cough and sore throat were the only clinical symptoms significantly associated with RSV detection. Among participants with RSV-only infection, 95.8% reported cough and 45.8% reported sore throat; corresponding values among those coinfected with influenza were 100% and 50.0%, respectively. This contrasted with significantly lower frequencies among virus-negative cases (71.5% and 25.0%, respectively), yielding p-values of 0.012 for cough and 0.009 for sore throat.

Comorbid conditions were disproportionately represented among coinfected individuals. Half of the RSV–influenza coinfected cases had documented asthma and heart disease, compared to <1% and 3.9% respectively among virus-negative participants. These associations were statistically significant (p < 0.001 for asthma, p = 0.018 for heart disease). Other conditions such as diabetes, hypertension, tuberculosis, and HIV did not differ significantly across infection categories.

### Factors associated with RSV Positivity

A multivariable Poisson regression model with robust variance was used to identify factors independently associated with RSV positivity among older adults. The analysis included 451 participants with complete data on all covariates (Table 4).

**Table 4.** Factors associated with RSV positivity among adults aged ≥65 years.

| **Risk factor** | **RSV prevalence % (95% CI)** | **Unadjusted PR (95% CI)\*** | **P value** | **Adjusted PR (95% CI)\*** | **P value** |
| --- | --- | --- | --- | --- | --- |
| **COVID period** |  |  |  |  |  |
| Pre-COVID | 7.1 (3.6, 13.8) | 1.00 |  | 1.00 |  |
| Post-COVID | 4.2 (2.7, 6.6) | 0.81 (0.33, 1.99) | 0.643 | 1.20 (0.44, 3.30) | 0.721 |
| **Season** |  |  |  |  |  |
| Dry | 5.3 (3.2, 8.8) | 1.00 |  | 1.00 |  |
| Wet | 4.3 (2.5, 7.5) | 0.85 (0.38, 1.89) | 0.690 | 0.96 (0.42, 2.18) | 0.923 |
| **Cough** |  |  |  |  |  |
| No | 0.7 (0.1, 5.1) | 1.00 |  | 1.00 |  |
| Yes | 6.2 (4.2, 9.1) | 7.18(0.98, 52.79) | 0.053 | 5.09(0.67, 38.43) | 0.114 |
| **Sore throat** |  |  |  |  |  |
| No | 3.1 (1.8, 5.5) | 1.00 |  | 1.00 |  |
| Yes | 7.9 (4.6, 13.7) | 2.24 (1.01, 4.95) | 0.046 | 1.66 (0.76, 3.60) | 0.202 |
| **Diarrhea** |  |  |  |  |  |
| No | 4.3 (2.8, 6.5) | 1.00 |  | 1.00 |  |
| Yes | 10.0(4.3,23.2) | 1.42 (0.44, 4.60) | 0.555 | 1.28 (0.37, 4.40) | 0.700 |
| **Heart disease** |  |  |  |  |  |
| No | 4.9 (3.3, 7.4) | 1.00 |  | 1.00 |  |
| Yes | 14.3(4.9,41.8) | 3.07 (0.99, 9.54) | 0.052 | 2.62 (0.88, 7.78) | 0.082 |
| **Asthma** |  |  |  |  |  |
| No | 5.0 (3.3, 7.4) | 1.00 |  | 1.00 |  |
| Yes | 50.0 (16.1, 99.9) | 6.79 (1.30, 35.46) | 0.023 | 6.08 (1.18, 31.26) | **0.031** |
| **Pneumonia** |  |  |  |  |  |
| No | 4.1 (2.7, 6.4) | 1.00 |  | 1.00 |  |
| Yes | 9.7 (4.5, 20.8) | 3.03 (1.26, 7.32) | 0.014 | 2.83 (1.06, 7.56) | **0.038** |

\*Based on complete data (N = 451). PR = prevalence ratio; CI = confidence interval.

In bivariate analysis, the presence of cough (PR: 7.18; 95% CI: 0.98–52.79; p = 0.053) and sore throat (PR: 2.24; 95% CI: 1.01–4.95; p = 0.046) were significantly associated with higher RSV period prevalence. However, after adjustment for covariates, neither symptom retained statistical significance.

Among comorbid conditions, asthma and clinician-diagnosed pneumonia were significant independent predictors of RSV infection. Individuals with asthma had a markedly higher period prevalence of RSV (aPR: 6.08; 95% CI: 1.18–31.26; p = 0.031), and those diagnosed with pneumonia during admission also showed increased period prevalence (aPR: 2.83; 95% CI: 1.06–7.56; p = 0.038). Heart disease showed a trend toward higher prevalence (aPR: 2.62; 95% CI: 0.88–7.78; p = 0.082) but did not reach statistical significance.

No significant differences in RSV period prevalence were observed by season (dry vs. wet), COVID period (pre-vs. post-pandemic), or gastrointestinal symptoms such as diarrhoea. The adjusted period prevalence in the wet season was comparable to the dry season (aPR: 0.96; 95% CI: 0.42–2.18; p = 0.923), and the post-COVID period was similar to pre-COVID (aPR: 1.20; 95% CI: 0.44–3.30; p = 0.721).

### Predictors of Severe RSV Illness (Hospitalization)

Hospitalization was used as a proxy indicator for severe RSV illness. Among 466 participants with complete data, we examined the association between selected comorbid conditions and hospitalization in RSV-positive cases (Table 5).

**Table 5.**
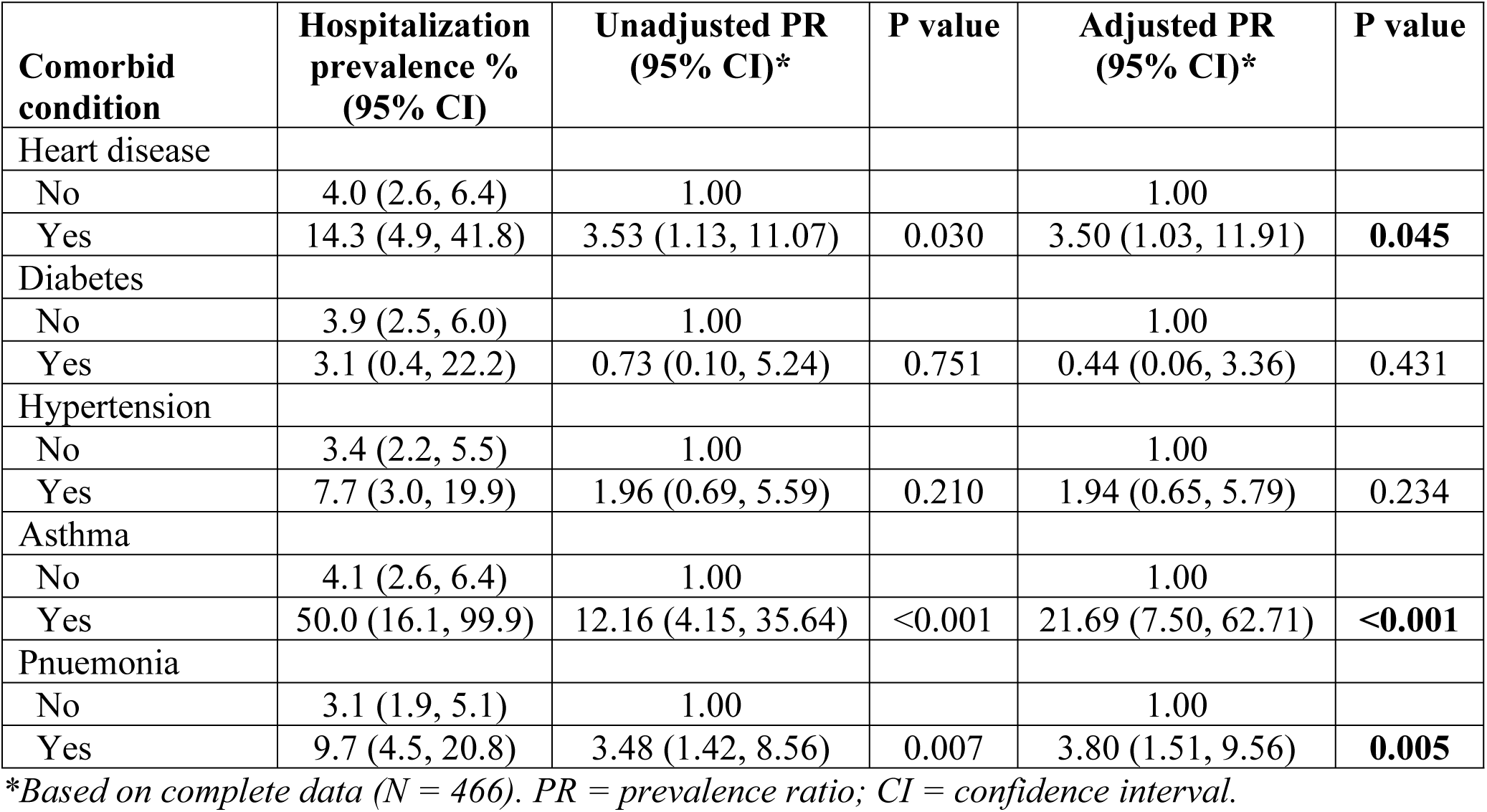
Comorbid conditions associated with hospitalization among RSV-positive adults aged ≥65 years.

Asthma was the strongest independent predictor of severe RSV illness. Half of RSV-infected individuals with asthma required hospitalization, compared to 4.1% among those without asthma. This corresponded to an adjusted prevalence ratio (aPR) of 21.69 (95% CI: 7.50–62.71; p < 0.001), indicating a more than twentyfold higher prevalence of severe illness.

Clinician-diagnosed pneumonia was also significantly associated with hospitalization, with an aPR of 3.80 (95% CI: 1.51–9.56; p = 0.005). Heart disease showed a similar pattern, with aPR = 3.50 (95% CI: 1.03–11.91; p = 0.045).

In contrast, neither hypertension nor diabetes showed significant associations with hospitalization in RSV-positive individuals. Hypertension was associated with a non-significant increase in hospitalization prevalence (aPR: 1.94; 95% CI: 0.65–5.79; p = 0.234), while diabetes had no meaningful association (aPR: 0.44; 95% CI: 0.06–3.36; p = 0.431).

## Discussion

Over 15 years of national ILI/SARI surveillance in Uganda, RSV emerged as an important contributor to respiratory disease among adults aged ≥65 years. Its period prevalence was lower than SARS-CoV-2, comparable to influenza A, and higher than influenza B, underscoring the need to recognize RSV alongside other major viral pathogens in this population. Unlike the typical symptom profile of acute respiratory infections, RSV was not strongly associated with common features such as cough or sore throat. Instead, it showed strong links with asthma and pneumonia, suggesting that RSV may act as a key underlying driver of clinical complications that often necessitate hospitalization. The circulation of RSV was also shaped by the COVID-19 pandemic: a marked suppression was observed during 2020–2021, coinciding with mitigation measures, followed by a clear re-emergence in 2022 and subsequent years.

### RSV prevalence in context

Pooled hospital-based data comprised of demographic and clinical characteristics over 15 years of sentinel surveillance indicate an RSV period prevalence of 4.8% among Ugandan adults aged ≥65 years, representing one of the few age-stratified estimates available from sub-Saharan Africa.. This estimate lies within the 3–7% annual attack rates reported for community-dwelling older adults in high-income countries (Falsey & Walsh, 2005; Kenmoe & Nair, 2024) and also comparable to hospital-based estimates in high-risk elderly subgroups with cardiopulmonary comorbidities (Falsey & Walsh, 2005; Havers et al., 2023). However from the East African Community, a systematic review and meta-analysis of acute respiratory infection studies reported a pooled RSV prevalence of approximately 11% (95% CI: 7–15%), with marked differences in country-specific rates (∼3% in Uganda, ∼6% in Kenya, and ∼29% in Tanzania), likely reflecting differences in climate, diagnostic methods, and study populations (Thérèse Umuhoza et al., 2021). Unlike much of the earlier regional data, which often relied on antigen-based assays and aggregated all ages, our study used systematic RT-PCR testing and applied strict ILI/SARI criteria to a defined elderly cohort across multiple ecological zones. The modest period prevalence observed here likely reflects both the restricted age range and the focus on medically attended illness, which captures more severe but fewer mild infections. Variability between our findings and pooled regional estimates highlights the importance of age-specific RSV surveillance to accurately characterise disease burden. Given Uganda’s growing elderly population and the availability of licensed RSV vaccines for older adults (Papi et al., 2023; Walsh et al., 2023), such data are critical to inform targeted prevention strategies, optimise clinical recognition, and integrate RSV testing into existing respiratory virus surveillance platforms.

### Clinical presentation

In our elderly cohort, cough and sore throat were the only symptoms significantly associated with RSV infection, although other respiratory and systemic features such as shortness of breath, fatigue, and myalgia were also observed. While cough was reported in over 95% of RSV-positive cases, it did not remain statistically significant at multivariable analysis, suggesting that although common, it may not be a reliable discriminator of RSV infection in this population.

This contrasts with reports from high-income settings where cough is considered a near-universal feature of adult RSV and is frequently accompanied by upper-respiratory symptoms such as sore throat and running nose rather than high fever (Falsey & Walsh, 2005; Sundaram, Meece, Sifakis, Gasser, & Belongia, 2014).

By contrast, fever could not be meaningfully evaluated as a distinguishing clinical feature in our cohort because it was a core component of both the ILI and SARI enrollment criteria. Its near-universal presence across all participants precluded assessment of its discriminative value for RSV. Importantly, this reliance on fever-based inclusion criteria may underestimate the true burden of RSV in older adults, as afebrile cases which are known to occur in this age group would have been excluded at the point of enrollment. Such afebrile presentations are biologically plausible, given the effects of immunosenescence. Age-related declines in innate and adaptive immune function blunt the production of pro-inflammatory cytokines and febrile responses, leading to attenuated systemic signs of infection (El Chakhtoura, Bonomo, & Jump, 2017).

Similar findings have been reported in LMIC-based surveillance studies, where reliance on fever-based case definitions has been shown to miss a substantial proportion of RSV cases among the elderly (Umuhoza et al., 2021). These observations underscore the need to revisit RSV case definitions for older adults and to incorporate clinical features beyond fever in order to improve case detection and surveillance accuracy.

### Role of comorbidities

Comorbidities are well-established modifiers of RSV susceptibility, clinical severity, and healthcare utilisation in older adults. In our cohort, asthma was the strongest independent predictor of RSV infection, with an adjusted prevalence ratio of approximately 20-fold. This very strong association is biologically plausible: chronic airway inflammation, structural remodelling, and impaired mucociliary clearance in asthma create a permissive environment for viral adherence, persistence, and enhanced symptom severity (Gay et al., 2024; Jesenak et al., 2023). Similar patterns have been observed instudies from North America and Asia, where asthma and chronic obstructive pulmonary disease (COPD) consistently emerge as major risk factors for RSV-related hospitalisation, prolonged recovery, and intensive care admission (Landi et al., 2024; Lee et al., 2013).

In this cohort, few RSV-positive cases were detected among patients with heart disease, likely reflecting under-reporting of comorbidities in routine sentinel surveillance data. Although the prevalence among cases with heart disease was modest, this remains notable given global evidence linking pre-existing cardiovascular disease to poorer RSV outcomes. Pathophysiologic mechanisms include reduced cardiopulmonary function, RSV-triggered myocardial stress, and systemic inflammation that can precipitate acute decompensation (Wyffels, Kariburyo, Gavart, Fleischhackl, & Yuce, 2020). These findings support the prioritizing of older adults with chronic respiratory or cardiovascular disease in RSV prevention strategies, including vaccination and early antiviral interventions where available. Strengthening comorbidity data capture in surveillance systems will improve risk stratification, while integrated care pathways may help mitigate the disproportionate RSV burden in these high-risk groups.

### Coinfection transmission patterns

In this elderly cohort, all viruses co-circulated year-round, but RSV–influenza coinfections were rare (0.4%), and no RSV–SARS-CoV-2 coinfections were detected. These findings are consistent with both high- and low-income country data, where dual RSV–SARS-CoV-2 infections remain infrequent and influenza–RSV co-detection in adults typically falls below 1% (Nickbakhsh et al., 2019; Stowe et al., 2021). Biologically, such transmission dynamics may be explained through innate immune response whereby by viral interference, whereby infection with one respiratory virus induces an antiviral state through type I and III interferon responses that suppress replication of other viruses (Laurie et al., 2015). Although innate immune responses may limit opportunities for viral co-infection (Rose et al., 2021), asynchronous epidemic peaks (particularly in East Africa, where RSV often peaks before influenza) can result in protracted outbreaks, with one virus dominating a season and another emerging sequentially.

Pandemic-era public health measures, including masking, physical distancing, and travel restrictions, likely further reduced simultaneous viral transmission, as observed in global surveillance reports (Olsen et al., 2021). Although the limited number of coinfections in our dataset precluded formal outcome comparisons, previous studies suggest that RSV monoinfection alone can cause substantial morbidity in older adults, with coinfection adding little or only modestly to clinical severity in this age group (Walsh et al., 2023).

Continued multi-pathogen testing in sentinel surveillance systems remains important to detect shifts in these patterns, particularly as COVID-19 control measures are relaxed and respiratory virus seasonality potentially re-aligns, increasing the theoretical window for coinfections.

### Seasonal and pandemic-related dynamics

RSV epidemics demonstrate marked geographic variation, shaped by the interplay of climate, viral ecology, and host susceptibility. In our dataset, peaks occurred consistently during May– June, coinciding with Uganda’s long rainy season; a pattern also observed in neighbouring Kenya (Rose et al., 2021). This contrasts with temperate regions, where RSV activity typically peaks in winter months, underscoring the influence of local climatic drivers on epidemic timing (Obando-Pacheco et al., 2018; Paynter, 2015). In tropical East Africa, high humidity may stabilise virions and facilitate transmission, while seasonal rainfall may increase indoor crowding, amplifying spread (Paynter, 2015).

The near-complete absence of RSV circulation in 2020–2021 parallels global reports of disrupted RSV seasonality during the COVID-19 pandemic, when non-pharmaceutical interventions (including masking, school closures, and mobility restrictions) suppressed respiratory virus transmission (Olsen et al., 2021). The subsequent resurgence in 2023 is consistent with the concept of “immunity debt,” in which reduced seasonal exposure temporarily increases population susceptibility, leading to atypical and often larger outbreaks once restrictions are lifted (Cohen et al., 2021; Delestrain et al., 2021).

These findings reinforce the importance of continuous, year-round RSV surveillance in LMICs to detect shifts in seasonal timing. As climate variability, urbanisation, and pandemic-related behavioural changes alter historical circulation patterns, predictive modelling and integration of RSV surveillance into early-warning systems will be critical for optimising vaccine deployment and strengthening clinical preparedness.

### Public Health and Clinical Implications

Our findings confirm RSV as a persistent contributor to acute respiratory illness and hospitalization among older Ugandan adults, despite its lower prevalence relative to SARS-CoV-2. By demonstrating strong associations with asthma and other chronic respiratory conditions, this study highlights the importance of prioritising high-risk groups (particularly older adults with chronic lung or cardiovascular disease) for preventive interventions once vaccines or monoclonal prophylaxis become available in LMICs.

For clinicians, heightened suspicion of RSV during seasonal peaks is essential, given its nonspecific presentation and overlap with influenza, COVID-19, and bacterial pneumonia. Incorporating RSV testing into diagnostic algorithms for older adults could improve case recognition, reduce inappropriate antibiotic use, and contribute to antimicrobial resistance mitigation. From a public health perspective, the seasonal concentration of cases suggests that future vaccination campaigns in Uganda could be optimally timed to precede the long rainy season. However, variability in climate and epidemic timing reinforces the need for sustained, year-round surveillance to guide implementation.

Beyond epidemic timing, continuous RSV surveillance will be critical for detecting post-pandemic shifts in circulation, monitoring the emergence of vaccine-escape variants, and informing cost-effectiveness models for preventive strategies. Integration of RSV testing into existing influenza and COVID-19 surveillance systems offers a pragmatic and resource-efficient path forward. Such measures would provide actionable evidence for policy, enhance preparedness for an aging population, and reduce the growing burden of respiratory morbidity in sub-Saharan Africa.

### Strengths and Limitations

This study has several notable strengths. It represents the most comprehensive analysis to date of RSV epidemiology among older adults in Uganda, and one of the few from sub-Saharan Africa. The 15-year period of continuous surveillance (2010–2025) enabled assessment of RSV patterns before, during, and after the COVID-19 pandemic, thereby capturing both long-term seasonal dynamics and pandemic-related disruptions. Inclusion of sentinel sites across diverse ecological zones—urban, peri-urban, and rural—enhanced representativeness for Uganda’s older adult population. Laboratory confirmation of RSV, influenza A/B, and SARS-CoV-2 using real-time RT-PCR ensured high diagnostic accuracy, while the collection of detailed demographic, clinical, and comorbidity data facilitated multivariable analyses of predictors of infection and hospitalization.

However, several limitations should be considered when interpreting these findings. First, the cross-sectional design, although based on long-term surveillance, limits causal inference regarding the temporal relationship between exposures (e.g., comorbidities) and RSV outcomes. Second, hospitalization was used as a proxy measure for severe disease; while pragmatic in a surveillance context, it may be influenced by health-seeking behavior, bed availability, and clinician admission practices, potentially leading to misclassification of severity. Third, comorbidity data relied on clinical records, which may underreport chronic conditions, particularly in settings with limited diagnostic capacity. Fourth, certain variables, including hospitalization status, had notable missingness, and although missingness was generally unrelated to RSV status, it could have reduced statistical power in some analyses. Fifth, the small number of RSV-positive cases with specific comorbidities (e.g., asthma) led to wide confidence intervals around some prevalence estimates, warranting cautious interpretation of effect sizes. Finally, the absence of virological subtyping and viral load data precluded exploration of potential differences in clinical presentation or severity by RSV subtype (A vs. B).

Despite these limitations, the study provides robust, nationally representative, and long-term evidence on RSV in older Ugandan adults. Future research should incorporate longitudinal cohort designs, improved comorbidity ascertainment, and molecular characterization of RSV strains to deepen understanding of clinical and epidemiological heterogeneity.

### Conclusions

This 15-year sentinel surveillance analysis is the most comprehensive evaluation of RSV in older adults in Uganda. RSV accounted for a consistent fraction of respiratory illness, comparable to influenza A, with a distinct seasonal peak during the rainy season. Asthma, pneumonia, and heart disease were key predictors of severe illness, while coinfections were rare. Pandemic-era suppression and later resurgence further illustrate the dynamic nature of RSV transmission. By filling an evidence gap in an underrepresented, high-risk population, this study supports integrating RSV into national surveillance frameworks, prioritising high-risk groups for prevention, and sustaining year-round monitoring to guide timely vaccine introduction.

## Data Availability

All relevant data are within the manuscript and its Supporting Information files.

## Acknowledgements

This work leveraged the Interdisciplinary Consortium for Epidemics Research (ICER) platform. We gratefully acknowledge the contributions of ICER in fostering collaboration across disciplines and providing critical support for advancing research on epidemic preparedness, response, and recovery.

